# Clinical Profiles of children born with Orofacial Clefts: Results from Fourteen East African Countries

**DOI:** 10.1101/2022.11.09.22282144

**Authors:** Fitsum Kifle Belachew, Desta Galcha Gerbu, Ermiyas Belay Weldesenbet, Eleleta Surafel Abay, Salome Maswime, Mekonen Eshete

## Abstract

**Background:** More than 100,000 cleft lip and palate patients have benefited from reconstructive surgeries in Africa because of surgical support from non-governmental organizations such as Smile Train. The Smile Train Express is the largest cleft-centered patient registry with over a million records of clinical records, globally. In this study, we reviewed data from the east African patient registry to analyze and understand the clinical profiles of cleft lip and palate patients operated at Smile Train partner hospitals in East Africa.

**Method:** A retrospective database review was conducted in all East African cleft lip and palate surgeries documented in the Smile Train database from November 2001 to November 2019.

**Results:** 86,683 patient records from 14 East African countries were included in this study. The mean age was 9.1 years, the mean weight was 20.2kg and 19kg for males and females, respectively, and 61.8% of the surgeries were performed on male patients. Left cleft lip only (n=22,548, 28.4 %) and right cleft lip only (n=17862, 22.5%) were the most common types of clefts, with bilateral cleft lip only (n= 5712, 7.2%) being the least frequent. Complete right cleft lip with complete right alveolus was the most frequent cleft combination observed (n = 16,385) and Cleft lip to cleft lip and palate to cleft palate ratio (CL:CLP: CP) was 6.7:3.3:1. Unilateral primary lip-nose repairs were the most common surgeries (69%). General anesthesia was used for 74.6 % (52847) of the procedures.

**Conclusion:** Most children with cleft lip and/or palate were underweight, possibly due to malnutrition or related to socioeconomic status. There were more male patients compared to females, which could be related to gender disparities. Access to surgical care for children born with congenital defects needs to be improved, and inequities need to be addressed via more evidence-based collaborative intervention strategies.

**Highlights:**

- More than 80,000 patient records from fourteen East African countries were analyzed in this study to describe orofacial clefts.
- Orofacial clefts were found most commonly in males, accounting for over 62% of all cases.
- To improve cleft care in East Africa, there is a greater need for evidence-based implementation of programs, research collaboration, and data-centric advocacy efforts.

## Background

Globally, 313 million major surgical procedures are performed each year. (1) Yet, unmet surgical needs persist in low- and middle-income settings such as Sub-Saharan Africa (SSA) (1). In this region, a great majority of the population is in the pediatric age group with 43 percent of the population being under the age of fifteen. The rate of surgical disease among the pediatric population is high, with the incidence of surgical disease accounting for 6-12 percent of pediatric admissions in SSA, and overall mortality of 12 percent (2). Existing evidence shows there is an association between unmet pediatric surgical needs and high rates of morbidity and mortality (3).

The Lancet Commission on Global Surgery, 2015 has collated compelling evidence on the state of surgery and strategies for improving access (4) The Commission had called for an increase in ‘‘demand-driven international surgical support aimed to address the unmet burden of surgical disease.’ In that regard, international non-governmental organizations (NGOs) have been playing a significant role in the delivery of surgical care in low and middle-income countries (LMICs), where there are huge disparities in access to timely surgical care. (5,6).

As part of its global efforts to improve access to pediatric surgical treatment, Smile Train (7), has made dedicated efforts in the last two decades toward improving access to cleft surgeries in LMICs. Cleft lip and palate (CLP) deformities have widely been neglected, despite having a significant impact on the lives of patients and their families. This is of particular concern in the SSA setup where the disease is associated with spiritual wrath and societal stigmas (8). The Smile Train and other collaborators’ initiative is bringing children out of the perceived veil of suppressed seclusion and educating communities that this is not a spiritual problem, but a correctable defect requiring surgical intervention.

Since its founding in 1999, Smile Train has been developing and implementing new, innovative approaches to delivering comprehensive cleft care (CCC) with the aim of enabling every child born with a cleft to live a full, productive life (9). To date, it has operated on more than 1.8 million cleft lip and palate patients in over 100 countries across the globe (10). This has resulted in possibly, the genesis of the world’s largest web-based data repository, the Smile Train Express (STX) with over a million patient records. These records have been made accessible to the organization’s partner hospitals for research and quality improvement initiatives (11).

Despite the open access to data, the clinical profile of patients undergoing cleft lip or palate surgery in SSA, particularly in East Africa, is poorly understood. In this study, our aim was to better understand the regional distribution of cleft surgeries, sociodemographic and clinical profiles of patients, surgical and anaesthesia techniques used, identify gaps, and recommend areas of improvement.

## Method

A retrospective descriptive study was conducted to analyze the clinical data in STX. Ethical approval was obtained from the Institutional Review Board (IRB) of Yekatit 12 Hospital Medical College in Addis Ababa under protocol number 60/20. Data was requested from the East African countries under study through a formal data-sharing agreement. 86,683 records spanning 20 years (November 2001 - November 2019).

East Africa is among the most populous subregions of Africa, with a population of approximately 455 million people across 18 countries. Most of this population lives in rural areas. In this region, Smile Train has long-standing surgical collaborations in 14 countries, operating in more than 40 hospitals. These countries, whose data are included in this study, include: Burundi, Djibouti, Ethiopia, Kenya, Madagascar, Malawi, Mozambique, Rwanda, Somalia, South Sudan, Tanzania, Uganda, Zambia, and Zimbabwe.

Once the data was obtained, it was exported into Stata version 16 statistical software (Stata Corp, College Station, TX, USA) for analysis. We used a visual and statistical approach to inspect for possible data encoding errors and data cleaning. We used descriptive and analytical methods to analyze the observed responses. For descriptive measures, we used frequency and percentage, while for categorical variables, we used the chi-square test of independence to evaluate any possible associations. A single sample proportion test was also utilized to see if the proportion of different types of clefts differed significantly from the prior trends. We used a p-value of 0.05 as significance for statistical decisions.

## Results

### Sociodemographic characteristics

A total of 86,683 patient records were included in this study from the Smile train East African partner countries. Ethiopia (41.8%, n= 36,253), Uganda (13.4%, n= 11,626), Kenya (13 %, n= 11,318), Tanzania (11.6%, n= 10,119), and Somalia (5.8%, n= 5,069) were the top five countries with the highest number of surgeries, with South Sudan (n=0.3%, 268) contributing the least to the regional pool of surgeries. 49,206 (61.8 %) of surgical patients were males and 30,306 (38.2 %) were females, while the remaining 1,171 were not clearly defined and stated with other unknown terminologies.

### Age distribution of study participants

As described in Table 1, 17451 (27.4%) of patients were aged less than or equal to one, and 63693 (39.3%) of patients were greater than seven years of age. The median and the interquartile range were 4 and 12, respectively, and the average age at presentation is 9.1 years.

### Type and distribution of cleft lips

The different types of orofacial clefts and their clinical and epidemiologic context were classified into the following: cleft lips (right and left), hard palate clefts (right and left), soft palate clefts, and alveolar clefts (right and left). These were further subclassified into “complete”, “incomplete”, “submucous” or “not applicable”. A significant number of responses (8.4 %, n = 7410) were not fit for analysis, referring to an error in the encoding or faulty entry of primary data, and so were excluded. As a result, the total (n) responses will vary for different variables in our reports.

The different types of clefts are listed in Table 2 below using the “LAHSHAL” phenotypic notation, due to its comprehensiveness and relatively high implementation rate globally. It makes use of mutually exclusive and inclusive categories - solitary cleft or in combination with other clefts in each category. With this analysis, the most frequent orofacial cleft is a unilateral cleft lip only, with a left cleft lip only (LCLO) incidence of 28.4% (n=22,548), followed by a right cleft lip only (RCLO) 22.5% (n=17,862), Bilateral Cleft Lip Only (BCLO) was found to account for the least occurrence in this regard at 7.2 % (5712). The most common combination of the cleft was found to be a complete right cleft lip with a right alveolus (n= 16,385) followed by a left lip with a left alveolar cleft (n=14,445). The least common occurrence in this category was an incomplete left cleft lip with an incomplete soft palate (n=160).

### Laterality

Furthermore, we used the above phenotypic description (listed in Table 2) of the clefts to summarize the various orofacial clefts accounting for laterality. Table 3 shows the counts of the various cleft types under each gender group as well as a p-value from the chi-square test of independence between the male and female sex.

The laterality of the orofacial clefts in 87.7 % (n=75,724) of patients was assessed by looking at the descriptions presented in Table 3. A 4.4:1 ratio laterality was found, with unilateral cleft patterns (n=61,261) outnumbering bilateral cleft (n=14,037).

A chi-square test of independence was used to determine the gender distribution of clefts. The test showed that the observed differences in the frequencies of clefts between males and females are statistically significant with a P-value of <0.001, except for patterns of the bilateral cleft lip only (P=0.426) only and the left cleft lip only (p=0.277).

A one-sided t-test revealed that females have a lower body weight (19kg) than males (20.2kg). Furthermore, a two-sample t-test indicated a statistical significance (p-value of 0.001) in gender differences in mean weight. When leveled in accordance using a simple age-based weight estimation method using the mean age of presentation (9.1 years), the estimated weight will be 36.4kgs. This shows a 16 kg difference, likely due to malnutrition considering the existing malnutrition burden in the region, and the feeding problems posed by the CLP anomaly itself.

### Surgical procedures

71% (n=61559) of patients in our sample pool had only one surgical procedure. It was difficult to confirm the type of surgery for 18% of the total sample, as the findings in the records were not valid. The remaining had multiple surgeries (two or more) and for purposes of simplicity, we did not list them in tabular format. Table 4 shows the surgical procedures and their distribution among male and female patients, indicating an obvious gender disparity in surgical interventions. General anesthesia was used for 74.6 % (52847) of the procedures, and local anesthesia for 15.3 % (10843). For 10.2 % (7171) of the procedures, the type of anesthesia was not registered.

## Discussion

With an analysis of more than 80,000 patient records, this study is one of the largest in not only the East African region but the continent at large. Our study revealed a mean age of 9.1 years at the first presentation for surgical intervention. This finding is consistent with earlier African studies, the cause of which has been described as a lack of community awareness, advocacy work/mobilization efforts, and poor birth defect detection and registry practices (12,13). This shows that whilst cleft is detectable in utero and at the time of birth, presentation for correction is still delayed on average to the age of 9 years in African communities. Though access to surgical care is rising, there is still a need for parental and caregiver education and health promotion programs outlining the benefits of early intervention (14–16).

The study has also revealed that a high number of patients were underweight at primary intervention, which could indicate an additional malnutrition burden. Although this finding may need to be supported by additional data and analysis, existing data indicate that CLP patients typically present with lower-than-expected weight, possibly owing largely due to feeding difficulties posed by the defect itself (18–21). As malnutrition is a bottleneck to timely surgical intervention, it is crucial to study its prevalence in such patients, and associated factors therein, to further support research-based efforts. In addition, poverty is a social determinant of health which may be linked to malnutrition, and poor access to surgical care.

A peculiar finding of this study is that a large majority of surgical treatments are performed on males (61.8%), and the cleft incidence is highly associated with males (p=0.001) - particularly cleft palate (CP) incidence (p=0.000). This finding supports the global data while at the same time differing from it and most other studies (13, 16, 22, 23). The gender disparities in the number of surgeries can be attributed to the gender preference exhibited by parents and or caretakers in bringing forth male children for treatment, as in many other developing countries (24,25).

In our use of phenotypic description (26), the LCLO (28.4%) and RCLO (22.5%) distributions go in line with earlier studies in Africa (22,27–29). Furthermore, the finding that complete cleft lips are more frequently associated with an alveolus cleft, is consistent with a previously acclaimed description (30).

The finding that BCLO as the least common orofacial (7.2%), differs significantly from earlier research conducted across Africa using Smile Train data (2007 - 2013), where cleft palate only (CPO) was the least prevalent type of orofacial cleft (16). On a related note, the CL:CLP:CP ratio in this study is 6.7:3.3:1. The cleft palate ratio is even lower than that of a previous study conducted in Africa, which indicated a ratio of 5.1:8.8:1 (13). This can be explained in light of the reality that the patients have higher rates of morbidity and mortality from poor feeding, and increased susceptibility to infectious diseases due to malnutrition, as described by other studies (13,31–33).

Most patients (69%) had a unilateral cleft lip repair, and the remaining procedures were performed in less than 12.2% of patients. This finding is consistent with the other study conducted on African content (13,34). The most frequently used anesthetic technique was general anesthesia (74.6%), a finding that goes in line with earlier research (34). General anesthesia has been previously proven to be the best mode of anesthesia for cleft lip surgery (35). However, recent trends indicate that the presence of underlying congenital anomalies in these patient groups, the risks associated with general anesthesia, and the anatomical challenges, local anesthetics were found to bring better perioperative outcomes (19,36–38). While the ultimate decision lies on the anesthesia care provider, the findings of this study, in correlation with existing data, are inclined towards the use of all practical anesthetic techniques and call for more research in this regard. Our findings further extend attention to the proper registration of anesthesia techniques, as a significant proportion of anesthesia provided to patients goes undocumented (10.2 %).

The findings of this study must be translated in light of the following limitations: it describes the patients at the time of admission to surgery and doesn’t include their postoperative clinical outcomes. As the postoperative period is critical in holistic clinical profiling, the authors concur with the notion that should data be available, it must be integrated in future analyses to devise comprehensive programs in the region.

## Conclusion and Recommendation

The key findings of the study are that children born with cleft lip and/or palate, present for primary correction at the mean age of 9 years, in addition, the majority of the children were underweighted possibly due to malnutrition, which could either be associated with feeding difficulties or related to their socioeconomic status. There were more male patients compared to females, which could be related to gender disparities and the social standing of girls in African communities. Health promotion and education in community programs can lead to earlier detection and intervention. Access to surgical care for babies born with congenital defects needs to be improved, and inequities need to be addressed via more evidence-based collaborative approaches. Furthermore, incorporating postoperative variables into the STX database is also important for having complete perioperative information on patients and for generating outcome data.

## Data Availability

Since it is patient data, Smile Train Inc. can provide the data upon request with appropriate ethical approval, but it is not publicly available.

## Authors’ Contributions

FK, DGG, EBW, ESA, and ME conceived and designed the paper. FK and EBW worked on data curation, formal analysis, and visualization. FK, DGG, and ME worked on data acquisitions and methodology. FK, DGG, and ESA drafted the manuscript. SM and ME supervised the research project. All authors critically revised and approved the final manuscript.

## Acknowledgment

Our grateful thanks go to Smile Train for providing the data we requested and for training DG, FK, and ESA at the Kenyan Medical Research Institute on research methodology.

## Supplementary files

**Figure 1:**
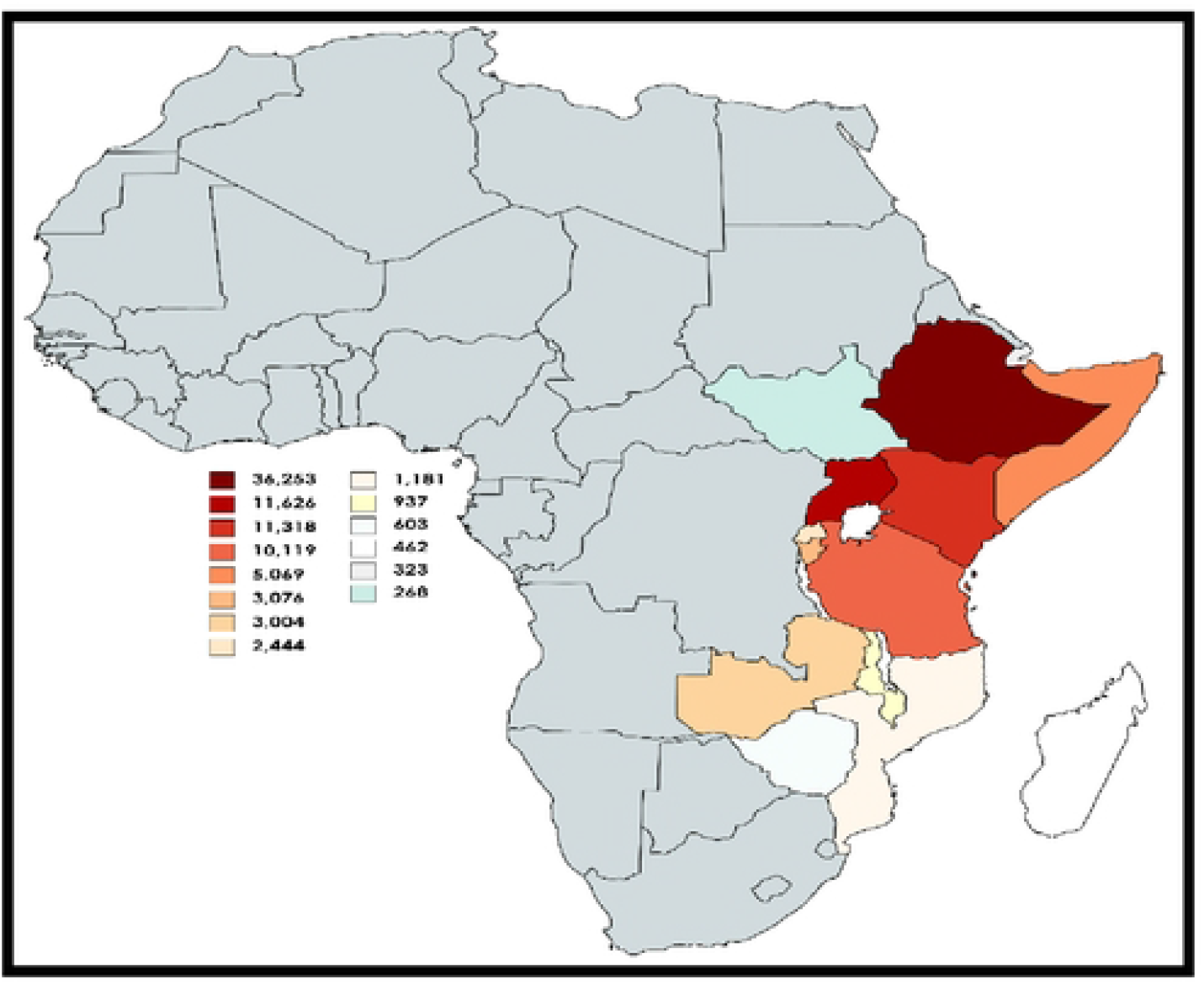
Top Contributing Countries

**Table 1:** Age distribution of study participants

| Age | Frequency | Cumulative frequency | Percentage |
| --- | --- | --- | --- |
| $\leq 1$ | 17527 | 17451 | 27.41 |
| $>1$ and $\leq 2$ | 7230 | 24646 | 11.31 |
| $>2$ and $\leq 3$ | 4110 | 28725 | 6.41 |
| $>3$ and $\leq 5$ | 5581 | 34279 | 8.73 |
| $>5$ and $\leq 7$ | 4368 | 38626 | 6.83 |
| $\geq 7$ | 25127 | 63693 | 39.3 |

**Table 2:** Phenotypic description of study participants

|  |  | *L* | *A* | *H* | *S* | *H* | *A* | *L* |
| --- | --- | --- | --- | --- | --- | --- | --- | --- |
|  | Response | Right cleft Lip | Right Alveolar cleft | Right Hard Palate | Midline Soft palate | Left Hard palate | Left Alveolus | Left Lip |
| Mutually Exclusive frequencies | Complete | 3164 | 37 | 293 | 804 | 721 | 171 | 3423 |
|  | Incomplete | 7694 | 18 | 119 | 196 | 1151 | 56 | 10110 |
|  | Submucous | - | - | 24 | 53 | 15 | - | - |
|  | Total | 10858 | 55 | 436 | 1053 | 1887 | 227 | 13533 |
| Not Mutually Exclusive total counts | Total | 28776 | 23066 | 18237 | 28072 | 21954 | 27316 | 27414 |
| Sum of mutual inclusive & exclusive counts | Total | 39,634 | 23,121 | 19,290 | 29,125 | 23,841 | 27,543 | 40,947 |

**Table 3:**
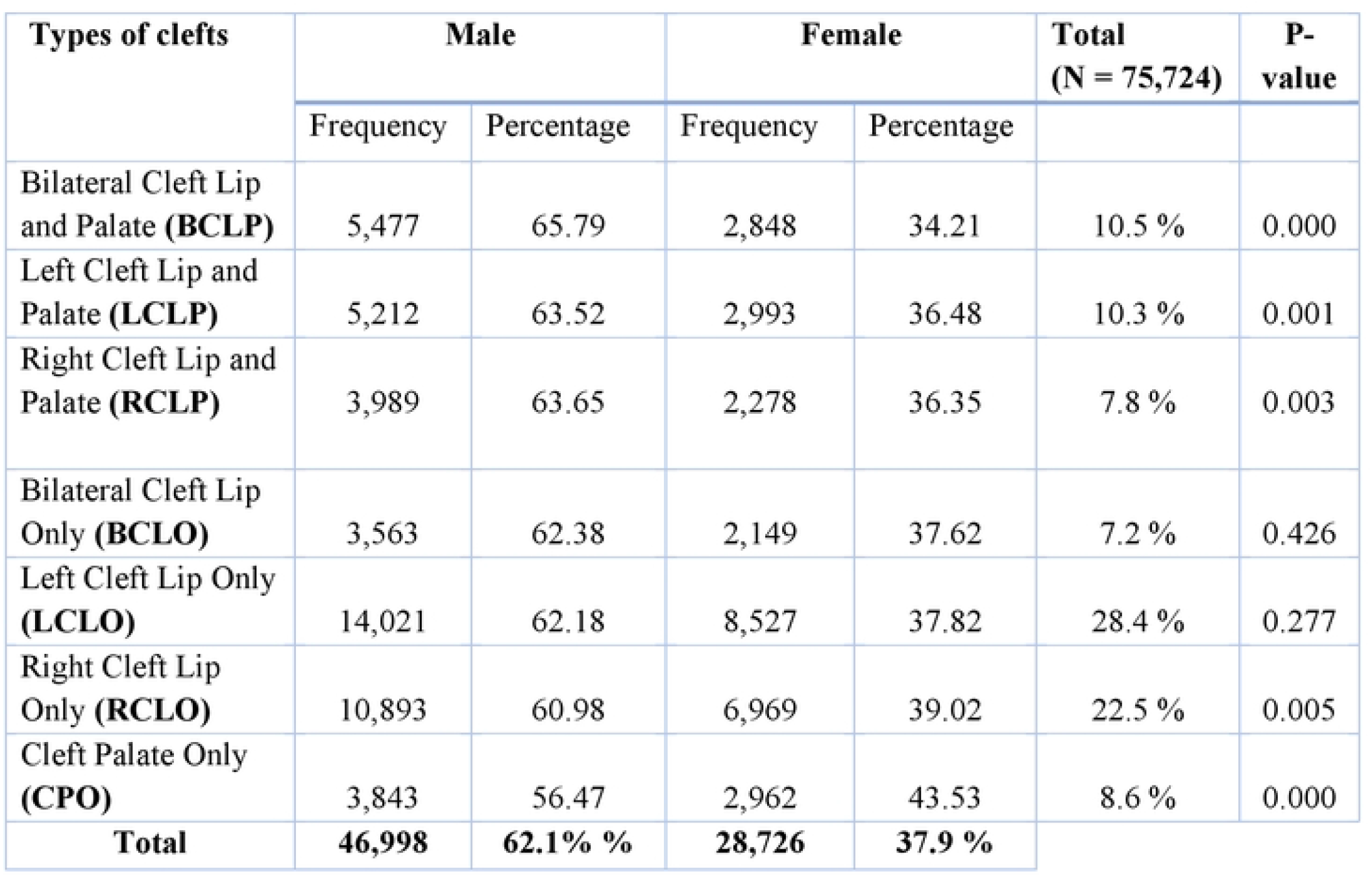
Types of orofacial clefts and their association with sex

**Table 4:** Surgical Procedures among males and females.

| Type of procedure | Male |  | Female |  | Total<br>n=61559 |
| --- | --- | --- | --- | --- | --- |
|  | Frequency | Percent | Frequency | Percent |  |
| Primary lip nose unilateral | 26430 | 62.1 | 16079 | 37.8 | 69 % |
| Primary lip nose bilateral | 4825 | 64.1 | 2696 | 35.8 | 12.2 % |
| Primary cleft palate | 4362 | 60.8 | 2805 | 39.1 | 11.6 % |
| Lip nose revision | 1516 | 58.1 | 1091 | 41.8 | 4.2 % |
| Fistula repair | 683 | 58.5 | 484 | 41.5 | 1.9 % |
| Secondary cleft palate | 208 | 56.5 | 160 | 43.4 | 0.6 % |
| Alveolar Bone Graft | 30 | 61.2 | 19 | 38.8 | 0.08 % |
| Other | 96 | 56.1 | 75 | 43.8 | 0.3 % |
| **Total** | **38150** | **62** | **23409** | **38** | **61559** |

## References

1. Rose J, Weiser TG, Hider P, Wilson L, Gruen RL, Bickler SW. Estimated need for surgery worldwide based on prevalence of diseases: a modelling strategy for the WHO Global Health Estimate. Lancet Glob Health [Internet]. 2015 Apr 27 [cited 2022 Jun 4];3 Suppl 2(Suppl 2):S13–20. Available from: https://pubmed.ncbi.nlm.nih.gov/25926315/

2. Debas HT, Donkor P, Gawande A, Jamison DT, Kruk ME, Mock CN. Essential Surgery. Disease Control Priorities, Third Edition (Volume 1): Essential Surgery [Internet]. 2015 Apr 2 [cited 2022 Jun 4]; Available from: https://www.ncbi.nlm.nih.gov/books/NBK333500/

3. Smith ER, Concepcion TL, Niemeier KJ, Ademuyiwa AO. Is Global Pediatric Surgery a Good Investment? World Journal of Surgery. 2019 Jun 30;43(6):1450–5.

4. Meara JG, Leather AJM, Hagander L, Alkire BC, Alonso N, Ameh EA, et al. Global Surgery 2030: evidence and solutions for achieving health, welfare, and economic development. International Journal of Obstetric Anesthesia [Internet]. 2016 Feb 1 [cited 2022 Jun 4];25:75–8. Available from: http://www.obstetanesthesia.com/article/S0959289X1500134X/fulltext

5. Patel PS, Chung KY, Kasrai L. Innovate Global Plastic and Reconstructive Surgery: Cleft Lip and Palate Charity Database. J Craniofac Surg [Internet]. 2018 Jun 1 [cited 2022 Jun 5];29(4):937–42. Available from: https://pubmed.ncbi.nlm.nih.gov/29485559/

6. Ng-Kamstra JS, Riesel JN, Arya S, Weston B, Kreutzer T, Meara JG, et al. Surgical Non-governmental Organizations: Global Surgery’s Unknown Nonprofit Sector. World J Surg [Internet]. 2016 Aug 1 [cited 2022 Jun 5];40(8):1823–41. Available from: https://pubmed.ncbi.nlm.nih.gov/27008646/

7. Ferry AM, Davis MJ, Rumprecht E, Nigro AL, Desai P, Hollier LH. Medical Documentation in Low- and Middle-income Countries: Lessons Learned from Implementing Specialized Charting Software. Plast Reconstr Surg Glob Open [Internet]. 2021 [cited 2022 Jun 5];9(6). Available from: https://pubmed.ncbi.nlm.nih.gov/34168942/

8. Kimotho SG, Macharia FN. Social stigma and cultural beliefs associated with cleft lip and/or palate: parental perceptions of their experience in Kenya. Humanities and Social Sciences Communications 2020 7:1 [Internet]. 2020 Dec 15 [cited 2022 Jun 5];7(1):1–9. Available from: https://www.nature.com/articles/s41599-020-00677-7

9. Volk AS, Davis MJ, Desai P, Hollier LH. The History and Mission of Smile Train, a Global Cleft Charity. Oral Maxillofac Surg Clin North Am [Internet]. 2020 Aug 1 [cited 2022 Jun 9];32(3):481–8. Available from: https://pubmed.ncbi.nlm.nih.gov/32471749/

10. Louis M, Dickey RM, Hollier LH. Smile Train: Making the Grade in Global Cleft Care. Craniomaxillofac Trauma Reconstr [Internet]. 2018 Mar [cited 2022 Jun 9];11(1):001–5. Available from: https://pubmed.ncbi.nlm.nih.gov/29387297/

11. Cleft Resources for Medical Professionals | Smile Train [Internet]. [cited 2022 Jun 4]. Available from: https://www.smiletrain.org/medical-professionals

12. Conway JC, Taub PJ, Kling R, Oberoi K, Doucette J, Jabs WW. Ten-year experience of more than 35,000 orofacial clefts in Africa. BMC Pediatr [Internet]. 2015 Feb 14 [cited 2022 Jun 30];15(1). Available from: https://pubmed.ncbi.nlm.nih.gov/25884320/

13. Conway JC, Taub PJ, Kling R, Oberoi K, Doucette J, Jabs WW. Ten-year experience of more than 35,000 orofacial clefts in Africa. BMC Psychiatry [Internet]. 2015 Feb 14 [cited 2022 Jul 7];15(1):1–9. Available from: https://bmcpediatr.biomedcentral.com/articles/10.1186/s12887-015-0328-5

14. Camille A, Evelyne AK, Martial AE, Denise K, Marie-Josée TA, Emmanuel K. Advantages of early management of facial clefts in Africa. International Journal of Pediatric Otorhinolaryngology. 2014 Mar 1;78(3):504–6.

15. Butali A, Mossey P. Epidemiology of Orofacial clefts in Africa: Methodological challenges in ascertainment. Pan African Medical Journal [Internet]. 2010 Feb 23 [cited 2022 Jul 6];2(1). Available from: https://www.ajol.info/index.php/pamj/article/view/51705

16. Azeez B, Keyla PR, Deborah VD, Ronald M, Mekonen AE, Wasiu LA, et al. Descriptive epidemiology of orofacial clefts in Africa using data from 46,502 Smile Train surgeries. Journal of Public Health and Epidemiology. 2017 May 30;9(5):114–21.

17. tinning K, Acworth J. Make your Best Guess: an updated method for paediatric weight estimation in emergencies. Emerg Med Australas [Internet]. 2007 Dec [cited 2022 Jul 7];19(6):528–34. Available from: https://pubmed.ncbi.nlm.nih.gov/18021105/

18. DSpace at My University: A study to assess the nutritional status of children with cleft lip and/or cleft palate and its correlation with breast feeding at a tertiary health care centre [Internet]. [cited 2022 Jul 7]. Available from: http://111.93.251.158/jspui/handle/123456789/1266

19. Sithole PA, Motshabi-Chakane P, Muteba MK. The characteristics and perioperative outcomes of children with orofacial clefts managed at an academic hospital in Johannesburg, South Africa. BMC Pediatrics [Internet]. 2022 Dec 1 [cited 2022 Jul 7];22(1):1–7. Available from: https://bmcpediatr.biomedcentral.com/articles/10.1186/s12887-022-03267-5

20. Escher PJ, Zavala H, Lee D, Roby BB, Chinnadurai S. Malnutrition as a Risk Factor in Cleft Lip and Palate Surgery. Laryngoscope [Internet]. 2021 Jun 1 [cited 2022 Jul 7];131(6):E2060–5. Available from: https://onlinelibrary.wiley.com/doi/full/10.1002/lary.29209

21. Chwa ES, Stoehr J, Gosain AK. 52. THE EFFECT OF PREOPERATIVE PATIENT CHARACTERISTICS ON POSTOPERATIVE OUTCOMES OF CLEFT PALATE REPAIR: AN ANALYSIS OF GLOBAL SMILE TRAIN DATA. Plastic and Reconstructive Surgery Global Open [Internet]. 2022 Apr 1 [cited 2022 Jul 7];10(4 Suppl):26–26. Available from: /pmc/articles/PMC8984344/

22. Hlongwa P, Levin J, Rispel LC. Epidemiology and clinical profile of individuals with cleft lip and palate utilising specialised academic treatment centres in South Africa. PLOS ONE [Internet]. 2019 May 1 [cited 2022 Jul 6];14(5):e0215931. Available from: https://journals.plos.org/plosone/article?id=10.1371/journal.pone.0215931

23. Yılmaz HN, Özbilen EÖ, Üstün T. The Prevalence of Cleft Lip and Palate Patients: A Single-Center Experience for 17 Years. Turkish Journal of Orthodontics [Internet]. 2019 [cited 2022 Jul 6];32(3):139. Available from: /pmc/articles/PMC6756567/

24. Swanson JW, Yao CA, Auslander A, Wipfli H, Nguyen THD, Hatcher K, et al. Patient Barriers to Accessing Surgical Cleft Care in Vietnam: A Multi-site, Cross-Sectional Outcomes Study. World Journal of Surgery 2017 41:6 [Internet]. 2017 Jan 24 [cited 2022 Jul 6];41(6):1435–46. Available from: https://link.springer.com/article/10.1007/s00268-017-3896-8

25. Poenaru D, Lin D, Corlew S. Economic Valuation of the Global Burden of Cleft Disease Averted by a Large Cleft Charity. World Journal of Surgery 2015 40:5 [Internet]. 2015 Dec 15 [cited 2022 Jul 6];40(5):1053–9. Available from: https://link.springer.com/article/10.1007/s00268-015-3367-z

26. Houkes R, Smit J, Mossey P, Don Griot P, Persson M, Neville A, et al. Classification Systems of Cleft Lip, Alveolus and Palate: Results of an International Survey: https://doi.org/101177/10556656211057368 [Internet]. 2021 Nov 23 [cited 2022 Jul 8]; Available from: https://journals.sagepub.com/doi/full/10.1177/10556656211057368

27. James O, Adekunle AA, Adamson OO, Agbogidi OF, Adeyemo WL, Butali A, et al. Management of Orofacial Cleft in Nigeria - A Retrospective Study. Annals of Maxillofacial Surgery [Internet]. 2020 Jul 1 [cited 2022 Jul 6];10(2):434. Available from: /pmc/articles/PMC7943976/

28. Moodley P, Sethusa MPS, Khan MI. The distribution of orofacial clefts at the medunsa oral health centre, cleft clinic. Journal of Cleft Lip Palate and Craniofacial Anomalies [Internet]. 2018 [cited 2022 Jul 6];5(1):20. Available from: https://www.jclpca.org/article.asp?issn=2348-2125;year=2018;volume=5;issue=1;spage=20;epage=27;aulast=Moodley

29. Bekele KK, Ekanem PE, Meberate B. Anatomical patterns of cleft lip and palate deformities among neonates in Mekelle, Tigray, Ethiopia; implication of environmental impact. BMC Pediatr [Internet]. 2019 Jul 24 [cited 2022 Jul 6];19(1):254. Available from: https://bmcpediatr.biomedcentral.com/articles/10.1186/s12887-019-1624-2

30. Geneser MK, Allareddy V. Cleft Lip and Palate. Pediatric Dentistry [Internet]. 2019 Jan 1 [cited 2022 Jul 9];77-87.e2. Available from: https://linkinghub.elsevier.com/retrieve/pii/B9780323608268000055

31. the Role of Artificial Eustachian Tube in Cleft Palate Patients | The Cleft Palate Journal [Internet]. [cited 2022 Jul 8]. Available from: http://cleftpalatejournal.pitt.edu/ojs/cleftpalate/article/view/99

32. Nirmala S, Saikrishna D. Dental concerns of children with lip cleft and palate - a review. Journal of Pediatrics & Neonatal Care [Internet]. 2018 Jul 19 [cited 2022 Jul 8];Volume 8(Issue 4). Available from: https://medcraveonline.com/JPNC/JPNC-08-00333.php

33. Smarius B, Loozen C, Manten W, Bekker M, Pistorius L, Breugem C. Accurate diagnosis of prenatal cleft lip/palate by understanding the embryology. World Journal of Methodology [Internet]. 2017 Sep 9 [cited 2022 Jul 9];7(3):93. Available from: /pmc/articles/PMC5618146/

34. Donkor P, Bankas DO, Agbenorku P, Plange-Rhule G, Ansah SK. Cleft lip and palate surgery in Kumasi, Ghana: 2001-2005. Journal of Craniofacial Surgery [Internet]. 2007 Nov [cited 2022 Jul 7];18(6):1376–9. Available from: https://journals.lww.com/jcraniofacialsurgery/Fulltext/2007/11000/Cleft_Lip_and_Palate_Surgery_in_Kumasi,_Ghana_.21.aspx

35. Law RC, de Klerk C. Anaesthesia for Cleft Lip and Palate Surgery. Medpharm [Internet]. 2014 [cited 2022 Jul 7];(14):27–30. Available from: https://www.tandfonline.com/doi/abs/10.1080/22201173.2007.10872500

36. Akitoye OA, Fakuade BO, Owobu TO, Efunkoya AA, Adebola AR, Ajike SO. Anaesthesia for cleft lip surgeries in a resource poor setting: techniques, outcome and safety. PAMJ 2018; 31:105 [Internet]. 2018 [cited 2022 Jul 7];31(105). Available from: https://www.panafrican-med-journal.com/content/article/31/105/full

37. Fontanals M, Merritt G, Sierra P, Echaniz G. Anesthetic Considerations and Complications of Cleft Palate Repairs. What’s New? Current Anesthesiology Reports 2021 11:3 [Internet]. 2021 Jul 1 [cited 2022 Jul 7];11(3):257–64. Available from: https://link.springer.com/article/10.1007/s40140-021-00460-7

38. Reena, Bandyopadhyay KH, Paul A. Postoperative analgesia for cleft lip and palate repair in children. Journal of Anaesthesiology, Clinical Pharmacology [Internet]. 2016 Jan 1 [cited 2022 Jul 8];32(1):5. Available from: /pmc/articles/PMC4784214/

